# Impact on mental health care and on mental health service users of the COVID-19 pandemic: a mixed methods survey of UK mental health care staff

**DOI:** 10.1101/2020.06.12.20129494

**Authors:** Sonia Johnson, Christian Dalton-Locke, Norha Vera San Juan, Una Foye, Sian Oram, Alexandra Papamichail, Sabine Landau, Rachel Rowan Olive, Tamar Jeynes, Prisha Shah, Luke Sheridan Rains, Brynmor Lloyd-Evans, Sarah Carr, Helen Killaspy, Steve Gillard, Alan Simpson

## Abstract

**Purpose:** The COVID-19 pandemic has potential to disrupt and burden the mental health care system, and to magnify inequalities experienced by mental health service users.

**Methods:** We investigated staff reports regarding the impact of the COVID-19 pandemic in its early weeks on mental health care and mental health service users in the UK using a mixed methods online survey. Recruitment channels included professional associations and networks, charities and social media. Quantitative findings were reported with descriptive statistics, and content analysis conducted for qualitative data.

**Results:** 2,180 staff from a range of sectors, professions and specialties participated. Immediate infection control concerns were highly salient for inpatient staff, new ways of working for community staff. Multiple rapid adaptations and innovations in response to the crisis were described, especially remote working. This was cautiously welcomed but found successful in only some clinical situations. Staff had specific concerns about many groups of service users, including people whose conditions are exacerbated by pandemic anxieties and social disruptions; people experiencing loneliness, domestic abuse and family conflict; those unable to understand and follow social distancing requirements; and those who cannot engage with remote care.

**Conclusion:** This overview of staff concerns and experiences in the early COVID-19 pandemic suggests directions for further research and service development: we suggest that how to combine infection control and a therapeutic environment in hospital, and how to achieve effective and targeted tele-health implementation in the community, should be priorities. The limitations of our convenience sample must be noted.

## Background

The WHO announced that COVID-19 infection was a pandemic on 11^th^ March 2020. Countries were advised to implement measures including social distancing, closure of schools and universities, home working and avoidance of travel. The potential impacts of the pandemic on people’s mental health and on the mental health care system are extensive. In the UK, a brief survey of members of the Royal College of Psychiatrists (RCPsych) in April 2020 predicted a ‘tsunami’ of demand in the coming weeks as people struggle to cope with COVID-19- associated social and economic stressors [1]. While many general population surveys have been launched, there has been less focus on the needs of people already living with mental health conditions, and on how mental health services are supporting them at a time of potential staff shortages and service reconfigurations. An editorial in *The Lancet Psychiatry* recognised this gap, arguing that “there has been far too little space dedicated to the status of those with severe mental illness who would usually receive community support, or on the problems faced on inpatient mental health units” [2].

Potential risks to provision of mental health care worldwide include staff absences due to sickness and the need to self-isolate, and workforce redeployment, for example from community to inpatient settings. In the community, staff in many countries have been required to limit face-to-face contacts to essential tasks such as the administration of injectable medication [3]. In the RCPsych survey, 43% of UK psychiatrists reported an increase in their urgent and emergency work and 45% a reduction in routine appointments [1]. Beyond the immediate changes to services seen in the early stages of the pandemic, there are many potential challenges that are specific to mental health care. These include difficulties in implementing infection control and social distancing guidance in settings where people may be very distressed or cognitively impaired [4], especially in mental health wards and the supported accommodation settings where many people with complex mental health problems live [5]. Face-to-face meetings are usually central to mental health care: severe restrictions to this seem likely to greatly alter staff and service user experiences. There is also a considerable risk that, even after restrictions are lifted, there will be a lasting exacerbation of health and social inequalities that affect people with longer-term mental health problems, for example, through increased economic disadvantage, inequalities in health care, or sequelae of increased trauma and abuse [6,7].

Since the start of the pandemic, experts from around the world have published views about potential negative impacts of the pandemic on mental health services [3,8,9] and the suggestion has also been recurrently made that it could provide an opportunity for positive service developments [10,11,12]. However, to date there has been no research directly assessing and reporting the experiences and perspectives of those currently working in the mental health system. Our aim was to inform further research and service responses by conducting, in the early stages of the COVID-19 pandemic, a survey of the perspectives and experiences of staff working in inpatient and community settings across the UK health and social care sectors.

## Methods

The King’s College London research ethics committee approved this study (MRA-19/20-18372), which involved mental health staff in the UK completing an online questionnaire.

### Study design

In the absence of a measure of pandemic impact on mental health care and mental health service users, we rapidly developed an online questionnaire to collect cross-sectional quantitative and qualitative data from mental health care staff.

### Participants

All staff working in face-to-face mental health care in the UK, or managing those who provide such care, were eligible to participate. All specialties were included, as were NHS, private healthcare, social care and voluntary sector services.

### Questionnaire: Development

The lead developer of the questionnaire, SJ, an academic and practising inner London psychiatrist, read key sources identified in an accompanying rapid review of relevant literature [13], including academic and professional journals, news media, and organisational websites, and followed relevant social media topics. The drafting of the questionnaire was further informed by the NIHR Mental Health Policy Research Unit (PRU) working group for this study (about 30 people, including clinicians, researchers and people with relevant lived experience), and the PRU Lived Experience Working Group. Both groups discussed the study at online meetings and identified important topics for inclusion. Nine further clinicians provided email summaries of the challenges they were currently facing and how they were being addressed.

Feedback was obtained from the PRU working group on a first draft of the questionnaire, together with additional input from experts in fields including mental health care for older people, children and adolescents, people with drug and alcohol problems, offenders, and people with intellectual disabilities. The questionnaire was revised and converted into an online format using the UCL Opinio platform. Pilot testing was then conducted with 17 clinicians who provided feedback on the length, acceptability and relevance of the questionnaire and on problems with specific items. Following this, a final version of the questionnaire was agreed.

### Questionnaire: Content

A mixture of structured and open-ended questions was included. Participants were asked which sector and region they worked in but not which organisation, maximising anonymity. Participants could skip questions if they wished, and internet cookies were used to prevent participants completing multiple questionnaires. A branching structure was adopted, with initial questions asking all participants to rate the relevance of each item on lists of:

- Challenges at work during the COVID-19 pandemic
- Problems currently faced by mental health service users and family carers (from a staff perspective)
- Sources of help at work in managing the impact of the pandemic

This was followed by sections for staff in specific settings and specialties. Questions also elicited details of adaptations and innovations introduced to manage the impact of the pandemic, and their perceived success, and enquired about concerns for the future and any aspects of current practice that they would like to keep after the pandemic. A copy of the survey is available at this web address: https://opinio.ucl.ac.uk/s?s=67819.

### Recruitment

Our aim was to achieve rapid recruitment of a large and varied sample by dissemination through multiple channels including:

- Professional networks, for example support from the Mental Health Nurse Academics UK, Unite in Health, Royal College of Psychiatrists and Royal College of Nurses.
- Social media, especially Twitter. Our partner, the Mental Elf, promoted the study frequently and advised us on social media strategy.
- Relevant mental health-focused bodies, including our partner the Centre for Mental Health and the Association of Mental Health Providers.

In the final week of recruitment, we targeted under-represented sectors, including relevant voluntary sector organisations and supported housing providers. We also sought to increase representation of staff from Black, Asian and Minority Ethnic groups by focused social media recruitment via the Mental Elf, including a video in which a prominent Black psychiatrist encouraged participation, and contact with the networks of PRU researchers who work on issues of diversity.

### Analysis

Quantitative data: Our main aim was to give an overview of the current impact of the pandemic: we produced descriptive statistics using Stata 15 to summarise relevant aspects of the quantitative data. Missing data are reported in the footnotes of the relevant tables.

Qualitative data: An analytical coding framework was developed guided by the study research questions, quantitative analysis results and themes emerging from initial survey responses. The responses to open-ended questions were left unedited and compiled into coding matrices in Microsoft Excel, with the emerging codes in the columns and cases in rows. Descriptive content analysis was then conducted, and all survey responses were indexed [14].

## Findings

We summarise key findings concisely here: our accompanying Supplementary report gives much more detail. Data were collected from 22 April 2020 to 12 May 2020. In total, 3,712 people started the survey (including many who clicked ‘Start’ but provided no or minimal data) and 1,793 got to the end. We report results for participants who completed at least one question from each of the three main sections open to all respondents. This produced a sample of 2,180. There were 15,010 responses to open-ended items, yielding 295,751 words for rapid qualitative content analysis.

### Participant characteristics

A large majority of participants worked in the NHS (1,935, 88.9%). Approximately a third described themselves as nurses (664, 30.6%), 347 as psychologists (16.0%), 254 as psychiatrists (11.7%), 97 as social workers (4.5%) and 80 as peer support workers (3.7%). Over a third identified as a manager or lead clinician in their service (826, 38.0%). Over two-thirds worked with working age adults (1,521, 70.0%), 39.2% worked with older adults (853), just under a third worked with people with learning disabilities (648, 29.8%), around a fifth worked with people with drug and alcohol problems (456, 21.0%) and another fifth worked with people with eating disorders (451, 20.7%). Participants could report working with multiple service user populations or in multiple settings. The majority worked in England (1,814, 83.4%) with around a third of those in England based in London (639, 35.3%) and a fifth in the North West (328, 18.1%). Four-fifths were female (1,378, 80.0%) and almost nine-tenths were from white ethnic groups (1,433, 87.0%). Full demographic details, including age, caring responsibilities and COVID-19 status, can be found in Table 1x of the Supplementary report (references to tables in the Supplementary report are herein indicated with an ‘x’ after the table number to distinguish them from tables in the main text).

**TABLE 1:**
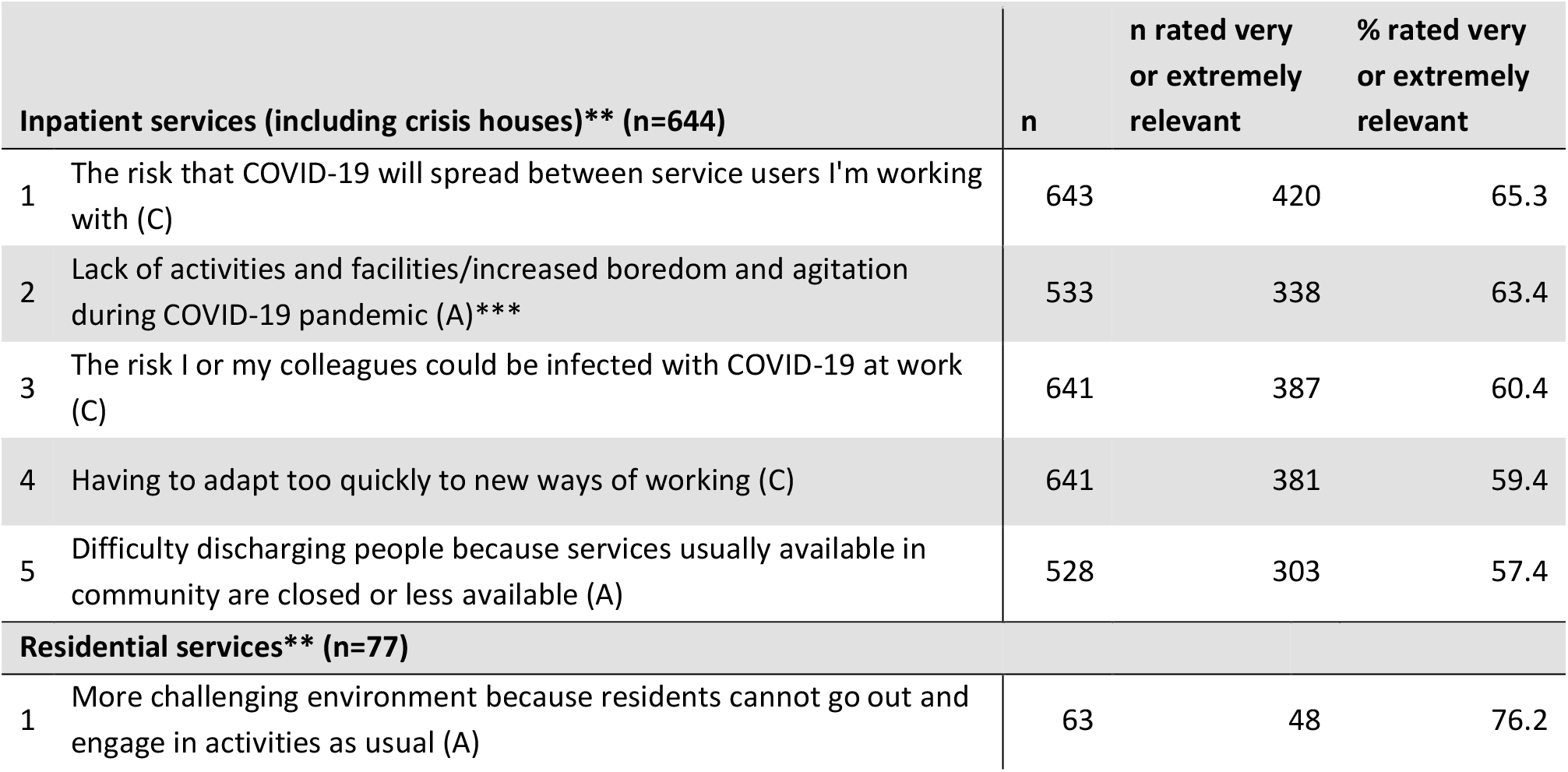
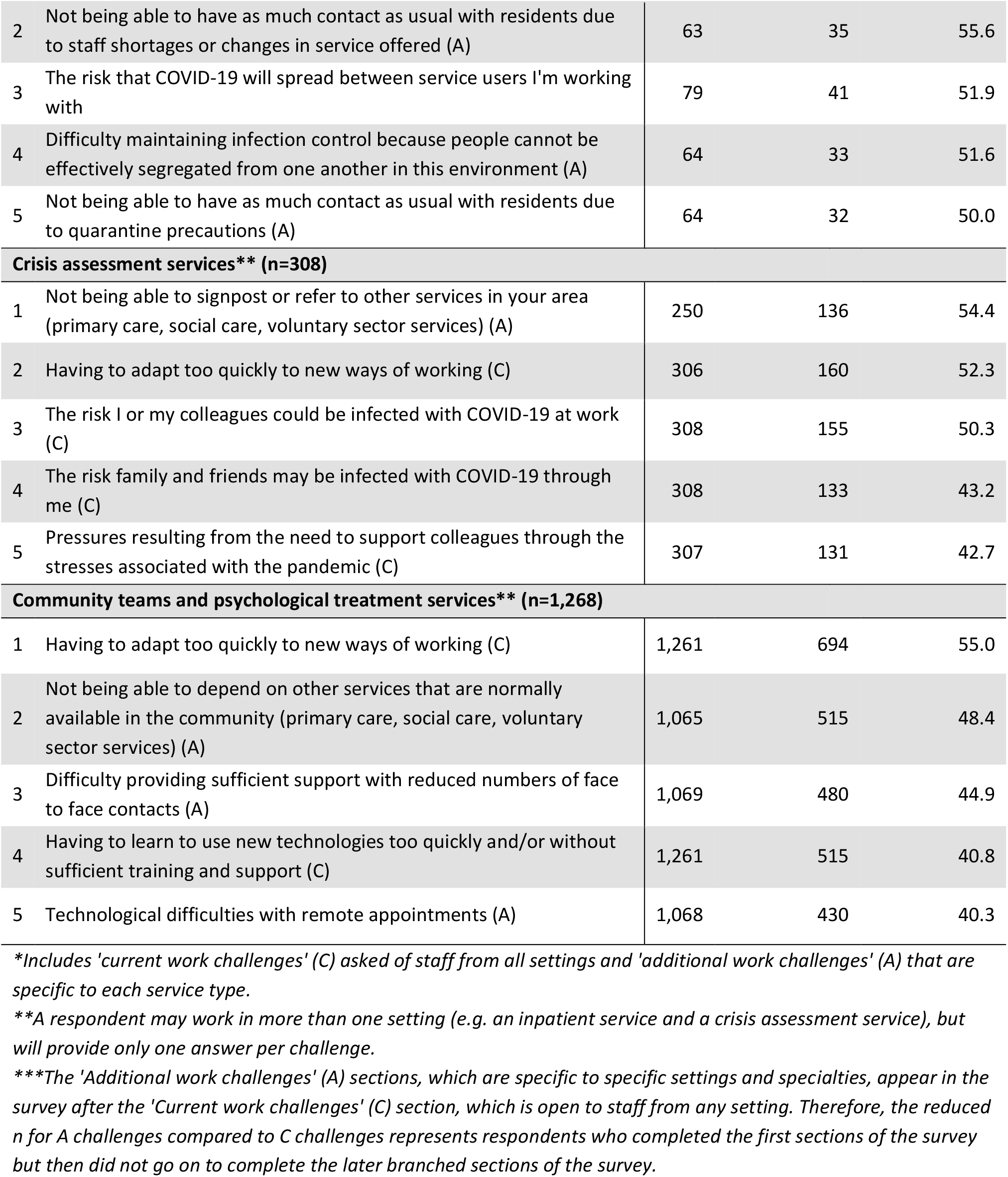
**Top five rated work challenges* for each setting *(See Tables 2x-6x and 29x-30x in the Supplementary report for further details)***

### Current challenges at work

Participants rated a list of current challenges at work, some general and others setting-specific, on a five-point scale from ‘Not relevant’ to ‘Extremely relevant’. Table 1 shows the five work challenges rated highest in each type of setting and Tables 2x–6x report this in further detail. In inpatient wards and crisis houses, infection control challenges, related to both service users and staff becoming infected, were rated highest, alongside increased boredom and agitation amongst service users due to lack of activity and contact on the ward. Crisis service staff rated as most relevant lack of services to which they could refer on or signpost. Community team staff rated items related to changes in ways of working and adoption of remote technologies highest, along with reduced availability of other services. The small group of residential service participants gave a high relevance rating to their environment being more challenging because residents cannot go out and/or engage in usual activities. Table 29x shows ratings by profession and Table 30x shows ratings by managerial roles. There were fewer obvious differences by profession than by setting, but managers and lead clinicians more often reported challenges relating to supporting colleagues with stressors due to the pandemic, and increased workload during the pandemic as very or extremely relevant (51.5% and 40.6%, respectively) compared to those not in these roles (31.8% and 21.3%, respectively).

**TABLE 2:** **Top five reasons infection control rules could not be followed for inpatient and community settings* (with frequencies), responses to an open-ended question *(See Tables 7x-8x in the Supplementary report for further details)***

| Inpatient and residential settings** | Community settings*** |
| --- | --- |
| 1 Conflict between infection control and providing care that is responsive and of good quality (98) | 1 Lack of space in office building for physical distancing (191) |
| 2 Service users who cannot or do not readily follow guidance (89) | 2 PPE availability (97) |
| 3 Guidance that conflicts or changes (76) | 3 Conflicts between infection control and providing care that is responsive and of good quality (72) |
| 4 Ward layout or office spaces that do not allow for social distancing (72) | 4 Facilities for using PPE, e.g. disposal, storage and removing of PPE (70) |
| 5 PPE availability (66) | 5 Impractical or inappropriate advice or guidance (54) |

\*A respondent may work in more than one setting (e.g. an inpatient service and a crisis assessment service).
\*\*Includes staff working in inpatient services, crisis houses and residential services.
\*\*\*Includes staff working in crisis assessment services, community teams and psychological treatment services, community groups and other settings.

### Impediments to infection control: Content analysis of qualitative responses

Half of staff in inpatient and residential settings reported they could not consistently follow the rules set on infection control (303, 50.5%), and just over a third reported they could not do this in community and other settings (518, 35.2%). Table 2 shows the impediments to this most often identified from qualitative content analysis of responses, with more detail in Tables 7x-8x. Tensions between meeting clinical needs and infection control were reported across settings, for example in responding to emergencies on wards or when service users in the community needed home visits, on which infection control measures were very difficult to implement. The built environment was the most frequently cited challenge in the community, and ward layouts impeded infection control in hospital. In each setting there were also reports of conflicting or unclear guidance. Reports of not having the facilities and processes to adhere to guidance, for example, in putting on and disposing of personal protective equipment (PPE), were especially prominent in the community. Unclear or conflicting guidance and procedures, and service users who are unable to understand and adhere to infection control rules, were reported across settings. Substantial numbers were also concerned about perceived conflicts between protective equipment and therapeutic relationships, for example, when trying to engage service users with paranoid ideas while wearing a mask.

### Service activity

We also asked participants to report, if data were available to them, the extent of activity change in the service in which they worked (Table 9x). Responses varied but reports of reduced activity considerably exceeded those of increased activity, especially regarding inpatient admissions (though less so for compulsory admissions) and new referrals to crisis services and community services. However, in community services, including psychological treatment services, similar numbers of staff said they were having more weekly contacts as said they were having fewer.

### Staff views regarding difficulties experienced by service users and carers with whom they are in contact

Table 3 summarises staff perceptions of the current relevance of various types of potential difficulty for the service users and carers with whom they were in contact (Table 10x reports this in greater detail and by service user group). Across all groups, staff tended to rate social difficulties as most relevant, for example, loneliness and a lack of usual support from family and friends. Several other types of problem were also rated by many staff as very or extremely relevant, including lack of normal support from mental health and other services, deterioration in mental health in the pandemic period, worries about infection and being at high risk if infected.

**TABLE 3:** **Summary of staff perspective on which of their service users’ and carers’ problems are most relevant, in order of % rated very or extremely relevant (n=2**,**180)** ***(See Table 10x in the Supplementary report for further details)***

|  | n | n rated very or extremely relevant | % rated very or extremely relevant |
| --- | --- | --- | --- |
| Lack of access to usual support networks of family and friends | 2,171 | 1,609 | 74.1 |
| Loneliness due to or made worse by social distancing, self-isolation and/or shielding | 2,180 | 1,504 | 69.0 |
| Lack of usual work and activities | 2,165 | 1,432 | 66.1 |
| Worries about getting COVID-19 infection | 2,166 | 1,334 | 61.6 |
| Lack of access to usual support from other services (primary care, social care, voluntary sector) | 2,173 | 1,333 | 61.3 |
| Worries about family getting COVID-19 infection | 2,169 | 1,296 | 59.8 |
| Increased difficulties for families/carers | 2,161 | 1,189 | 55.0 |
| Lack of access to usual support from NHS mental health services | 2,172 | 1,120 | 51.6 |
| Relapse and deterioration in mental health triggered by COVID-19 stresses | 2,180 | 1,010 | 46.3 |
| High personal risk of severe consequences of COVID-19 infection (e.g. due to physical health comorbidities) | 2,168 | 988 | 45.6 |
| Increase in reliance on family/family tensions | 2,164 | 892 | 41.2 |
| Difficulty understanding or following current government requirements on social distancing, self-isolation and/or shielding | 2,168 | 882 | 40.7 |
| Difficulty engaging with remote appointments by phone or via digital platforms | 2,172 | 862 | 39.7 |
| Increased risk from abusive domestic relationships | 2,165 | 821 | 37.9 |
| Diminished access to physical health care for problems other than COVID-19 | 2,172 | 735 | 33.8 |
| Having to stay at home in poor circumstances, or not having a home to go to | 2,169 | 752 | 34.7 |
| Difficulty getting food, money or other basic resources | 2,162 | 714 | 33.0 |
| Effects of COVID-19-related trauma | 2,165 | 597 | 27.6 |
| Risk of increased drug and alcohol use or gambling | 2,167 | 595 | 27.5 |
| Lack of access to or of equitable provision of physical healthcare for COVID-19 | 2,173 | 504 | 23.2 |
| Loss of liberty and rights due to changes in implementation of mental health legislation | 2,165 | 412 | 19.0 |
| Lack of access to medication and to processes for administering and monitoring it | 2,172 | 383 | 17.6 |
| Problems with police or other authorities because of lack of understanding of /ability to stick to current government requirements | 2,170 | 295 | 13.6 |

Responding to open-ended questions, staff identified a range of groups of service users about whom they were currently particularly concerned, some because of impacts on their clinical condition, others because of their social characteristics or circumstances, or because of specific difficulties providing an adequate service for them. Table 4 summarises groups frequently identified as of particular concern, and Supplementary Table 11x gives more detail.

**TABLE 4:**
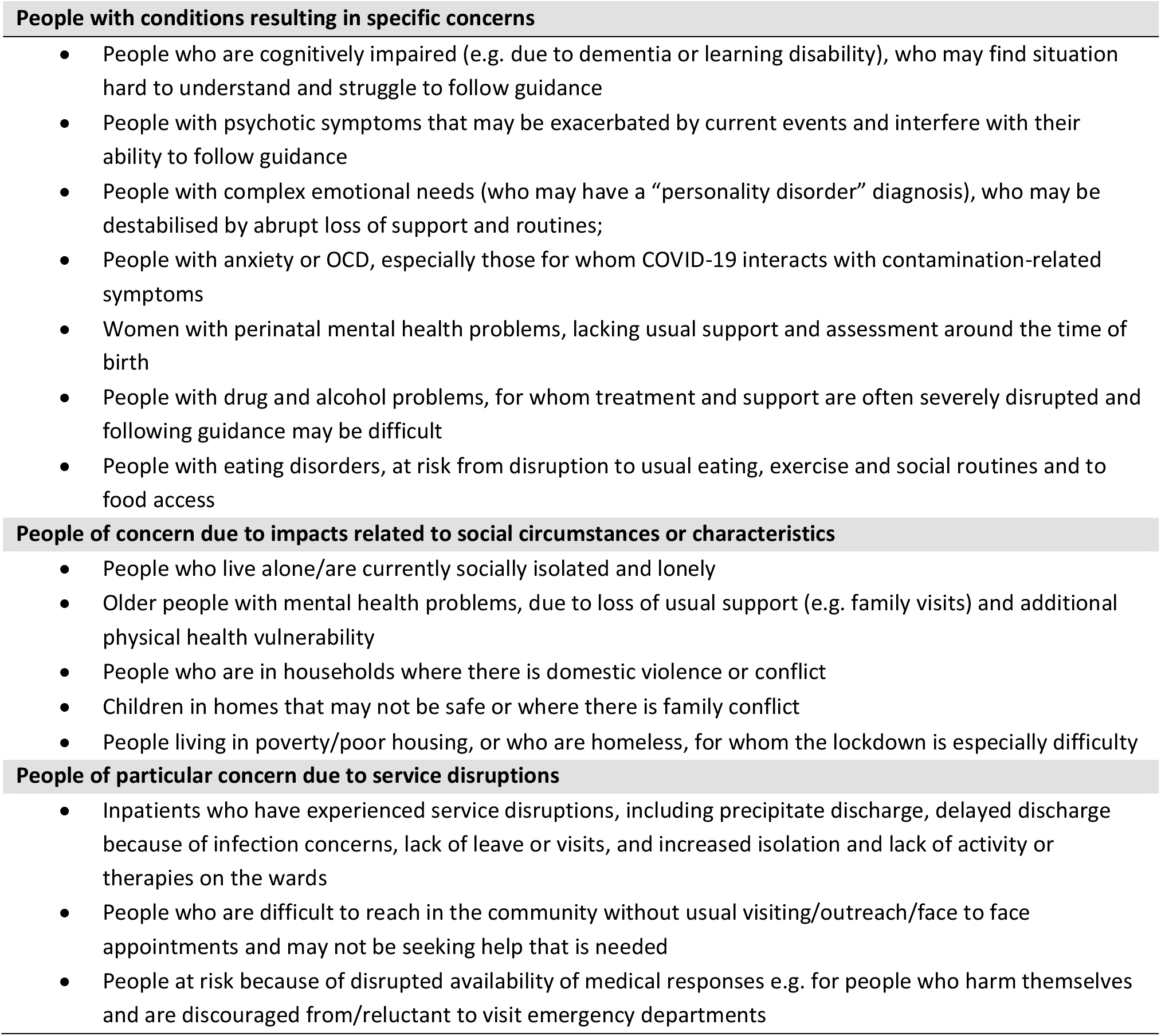
**Frequently cited examples of the groups of service users about whom staff participants have been especially concerned during the pandemic: qualitative content analysis of open-ended responses** ***(See Table 11x in the Supplementary report for further details)***

We also asked staff whether they were seeing people with mental health difficulties that appeared to arise from the pandemic (Table 12x). Some described symptoms directly related to COVID-19, such as delusional beliefs regarding COVID-19 infection or quarantine, and health anxiety or obsessive-compulsive symptoms related to infection. Others described relapses in people who had long been stable that they felt were linked to the stresses of the crisis. Some also reported apparently first presentations of mental health problems such as psychosis or mania among healthcare workers.

### Sources of help at work

Table 5 summarises responses to a question about which sources of help were currently most important to staff in managing the impact of COVID-19 at work. Across all professions, the most important sources of help were support and advice from employers, colleagues and managers, closely followed by new digital ways of working and the resilience and coping skills of service users and carers, the latter presumably seen as making the crisis less burdensome for staff, at least at its onset. Patterns of response were not markedly different across professional groups (Tables 13x-14x).

**TABLE 5:** **Summary of sources of help in managing COVID-19 impacts at work, in order of % rated very or extremely important (n=2**,**180)** ***(See Tables 13x-14x in the Supplementary report for further details)***

|  | n | n rated very or extremely important | % rated very or extremely important |
| --- | --- | --- | --- |
| Guidance from my employer on managing clinical and safety needs due to COVID-19 | 2,180 | 1,415 | 64.9 |
| Support and information from colleagues | 2,172 | 1,405 | 64.7 |
| Support and advice from my manager(s) | 2,169 | 1,374 | 63.3 |
| Adoption of new digital ways of working | 2,165 | 1,322 | 61.1 |
| Resilience and resourcefulness in adversity among service users and carers | 2,180 | 1,309 | 60.1 |
| Guidance disseminated by the NHS or professional bodies | 2,167 | 1,277 | 58.9 |
| Being aware of public support for key workers | 2,165 | 963 | 44.5 |
| Staff well-being initiatives set up during COVID-19 in my workplace | 2,160 | 855 | 39.6 |
| National initiatives to support service users and carers, such as helplines and online peer support | 2,155 | 823 | 38.2 |
| New initiatives in NHS mental health services | 2,158 | 815 | 37.8 |
| The support offered by local volunteers and mutual aid groups | 2,166 | 783 | 36.2 |
| Support and new initiatives from local voluntary sector organisations | 2,161 | 763 | 35.3 |
| National initiatives to support staff well-being | 2,156 | 714 | 33.1 |
| Information from the media or social media | 2,169 | 472 | 21.8 |

### Service changes and adaptations

Participants in crisis and community services were asked whether services they worked in had changed opening hours or locations, and how their practices had changed (Table 15x). Services that had increased their hours during the crisis, for example with weekend opening, were described, as well as reductions in other services. Most staff working in crisis services reported that home visits were continuing when strictly necessary. A mixture of responses was obtained from community services (including both community mental health teams and psychological treatment services), with some reporting continuing face-to-face contacts and home visits as needed, others having stopped them. Responses regarding psychological treatment were split between aiming to provide a full treatment by video call or phone, and providing abbreviated contacts only.

Open-ended questions elicited reports of adaptations and innovations made to manage the impact of the pandemic (Table 16x). The most widely reported shift was greatly increased adoption of remote technologies, as discussed below. Some participants also reported adopting new digital tools for assessment and therapy, such as apps and websites. Other innovations included new crisis services, such as crisis assessment centres rapidly established as alternatives to hospital emergency departments and new crisis phone lines, and re-organised services, resulting in extended hours, increased access for specific groups or shorter waiting lists (e.g. for psychological treatment). Reported changes in the types of help offered included community services arranging practical help, such as food deliveries for service users, and providing resource packs to help service users to be active at home. Also frequently described were new or expanded forms of support for staff, including ‘wobble’ rooms (quiet rooms for staff who feel overwhelmed), staff helplines, increased supervision, wellness check-ins, and more use of informal support mechanisms.

Also reported was a general shift towards a more flexible approach, reducing bureaucracy and removing barriers to change, leading to a more agile way of working and a more responsive service. Many staff also valued the many benefits to their wellbeing, productivity and efficiency in being able to conduct some of their client contact or administrative tasks away from the office.

### What’s working and what’s not in remote working: qualitative analysis of open-ended responses

Further quantitative and open-ended questions explored views and experiences of the shift to remote working (Tables 17x-19x). Almost all staff in community services (1,011, 94.1%), and a large majority in crisis services (219, 83.0%), were replacing some face-to-face contacts with phone or video calls. The shift to video calls did not appear to have been very extensive, however, with the majority (475, 54.5%) reporting use of this technology for 20% or fewer of their patient contacts.

Views about this were mixed. Video calls for communication between staff attracted the greatest enthusiasm, with more than two-thirds (815, 73.4%) from both community and crisis services agreeing or strongly agreeing that they are a good way to hold staff meetings; this was echoed in open-ended questions. A majority (818, 74.0% of respondents to this question) agreed or strongly agreed that video calls were a good way to assess progress of some people already known to the service, but only 39.8% (442) agreed or strongly agreed that they can be a good way of making initial assessments.

Responses to open-ended questions (Table 6, Tables 18x-19x) likewise identified concerns about being able to make a good assessment remotely, as well as about forming rapport: they tended to suggest digital technologies were useful for clients with less complex needs, for “light-touch” interventions or for low intensity therapeutic approaches and follow-up appointments. A majority (725, 65.8%) agreed or strongly agreed that use of remote rather than face-to-face consultations had resulted in not having contact with some service users who had not engaged with remote appointments. Other impediments to remote working, identified through open-ended questions (Table 19x), included accessibility issues for service users with cognitive impairments or dementia, or who lack access to appropriate technology, relevant skills or privacy at home, and outdated or scarce IT equipment within services.

**TABLE 6:** **Key points on remote working – what’s working well and what’s not** ***(See Tables 17x-19x in the Supplementary report for further details)***

| What's working well in tele-health | What can prevent tele-health from working |
| --- | --- |
| • **Efficiency of remote working:**<br>○ Allows prompt responses<br>○ Saves travelling time<br>○ Is better for the environment<br>○ May be more convenient for both staff and service users<br>○ Allows staff to connect easily with each other, even if based in different places and different teams<br>○ Allows home working<br>• **Best alternative for now:**<br>○ Remote working is allowing services to keep going despite infection control restrictions<br>○ Innovative use of IT and digital tools can allow group programmes or individual therapies to continue successfully<br>• **Benefits for some clients:**<br>○ some clients are happy with video-call technology and even prefer it<br>○ Access is improved for some people, especially if travel and public places are challenging<br>○ May be an efficient way of helping people with less complex needs | • **Inadequate resources:**<br>○ Equipment and internet connections of low quality<br>○ Processes and preferred platforms not clearly established.<br>○ Staff may lack training and confidence<br>• **Impacts on communication and therapeutic relationships**<br>○ May be harder to establish and maintain a good therapeutic relationship<br>○ May be harder to make an assessment, especially at first contact<br>○ May be challenging for longer, more in-depth sessions<br>• **Digital exclusion:**<br>○ People who lack equipment and resources to connect<br>○ People who don't have skills or confidence to connect (including people with cognitive impairments)<br>○ People lacking a suitably private environment for remote appointments<br>• **Service user preferences:**<br>○ Some service users strongly prefer confidential conversations to be face-to-face, or may feel suspicious or anxious about remote means<br>○ If they do accept remote contacts, some prefer simpler phone or messaging modalities<br>○ Some service users do not engage with remote contacts |

### Future hopes and concerns

67.8% answered yes to a question asking whether they wished to retain longer term any of the changes made during the pandemic. Table 20x summarises qualitative analysis of responses. A large majority involved keeping some aspects of remote working, with many feeling that selective use of technology platforms to connect staff with each other and with service users has potential long-term benefits for efficiency and the environment, particularly if technical difficulties are resolved and appropriate protocols developed. Others wished to retain some new service initiatives, such as crisis centres in the community, or the increased flexibility and ease of making changes experienced at this time.

Responses to a question about concerns for the future were numerous and detailed (Table 21x). While many participants reported that referrals to their service had decreased in the early phase of the pandemic, many feared that need would increase significantly in future and that lack of capacity and staff burnout may impede response to this. Anticipated drivers of increased future need included traumas, bereavement and complex grief experienced by frontline staff, service users and the wider public; mental health problems not managed effectively among people who have disengaged or not sought help during the pandemic; increased levels of domestic abuse and family conflict; and the effects of wider societal disruption and increased inequalities due, for example, to unemployment and homelessness. Fears were also expressed that reduced levels of service might persist inappropriately after the current emergency period, that changes made in response to the crisis might be used to justify reduced funding in future, or that staff would be expected to continue with working patterns that they had agreed to only because of the crisis. Extension of remote working beyond the circumstances in which it had proved helpful was a further concern. Several respondents were concerned about the disproportionate impact of the pandemic on Black, Asian and Minority Ethnic staff and service users, and about potentially increased racism and xenophobia.

## Discussion

A wide range of challenges are being experienced across the mental health sector, some specific to service settings or groups of service users and carers. While many commentators have predicted a significant and widespread impact of COVID-19, we are able to provide a more detailed report that is rooted in direct experience of the effects of the pandemic on mental health care, albeit only in one country and only from the perspective of practitioners.

In the context of the pandemic, infection control is an immediate need whose complexity in mental health settings is a significant finding from our study. Lack of PPE was sometimes identified as a problem. More prominent, however, were challenges relating to processes, to the physical environment in which mental health care is delivered, and to tensions between infection control requirements and providing safe care and maintaining therapeutic relationships with people who may be distressed, suspicious or struggling to comprehend the situation. Inpatient and residential services, and crisis services, where continuing face-to-face contacts appear more frequent than in routine care, are not surprisingly the settings in which staff are most immediately concerned with the spread of infection: the price of failure is potentially very high, as indicated by a recent Care Quality Commission report on excess deaths related to COVID-19 among people subject to the Mental Health Act [15].

The shift to remote working, strikingly rapid given that telehealth has been discussed over many years but with limited implementation, has been widely discussed; we examine staff perspectives on this in detail in the current study. Both our quantitative and qualitative data suggest clear support for its partial adoption in the longer term: remote contacts are seen as valuable for staff meetings, and for convenient and environmentally friendly follow-up of well-engaged clients with access to and a positive view of technology. However, staff give a very clear warning that there are still important technological, social and procedural barriers to be addressed, and that its use should remain selective, complementing rather than replacing face-to-face contact.

This and other innovations that we document above suggest that, as in other domains of healthcare, there has been considerable agility and flexibility in at least some service contexts during the current crisis, with urgent needs overcoming well-documented barriers to implementing new ways of working. However, while responses to our question about innovations that staff would like to retain were numerous, serious concerns regarding both the short and long-term future were also widely expressed: these data were collected at a very early stage in the COVID-19 pandemic.

### Limitations

This is only one perspective on the impact of the pandemic on mental health care, albeit one rooted in direct experience: it will be essential to investigate service user and carer perspectives, and to measure impacts on the mental health system more systematically as further data become available. Given the unprecedented pace of change in the world and in mental health services, we prioritised gaining a broad overview of impacts and responses, but much detail will have been missed. Our questionnaire was by necessity an *ad hoc* and not an established and validated tool. Our sample, gathered by disseminating our questionnaire through a range of channels, is not representative of those who work in mental health care settings, and may either over-represent people who have strong concerns about the situation or those who wish to report successful new practices. We managed to include a range of professions and work settings but did not recruit as successfully as we had hoped outside the NHS – more targeted efforts and time are likely to be needed to reach relevant staff from other sectors. Many people with mental health difficulties also come into contact with GPs, pharmacists, paramedics, and A&E doctors and nurses, especially if they are not under secondary services; we have not included these perspectives. We are especially concerned that, while we do not have any definitive overall figure for the UK mental health care workforce, it is clear that the number of non-White participants in our survey is relatively small, despite targeted efforts to increase their number and a strong emphasis on anonymity and confidentiality, as advised in previous discussions of this frequently experienced difficulty [16]. Further efforts to engage and form partnerships are likely to be needed here too.

### Implications

We present here a series of snapshots capturing, from a staff perspective, the situation in mental health care services in the rapidly evolving early stages of the COVID-19 pandemic. This work cannot yield definitive answers and should be interpreted alongside other perspectives, but offers researchers, services and policymakers directions for service development and further rapid research. Regarding immediate priorities, our findings point to specific challenges to be addressed to achieve more successful infection control. Remote working is a further immediate focus for research and service developments. Participants’ accounts suggest that it has been helpful in keeping services going and maintaining some level of contact in the community, and for communication between staff, but that there is now a need to develop clearer processes in collaboration with service users for its targeted use, to implement guidance and evidence that already exists, and to explore ways of overcoming barriers to its effective use. Finally, we suggest that our findings are a mixture of the predictable and the unpredictable, for example, in relation to levels of service activity. They indicate that it will be essential to continue to observe carefully what is occurring in mental health care settings around the world, tailoring responses to real needs and pressures, not to predictions and expectations.

## Lived experience commentary: Rachel Rowan Olive and Tamar Jeynes

It is reassuring to see that staff share many of our concerns about the Covid-19 pandemic: premature discharges, isolation, difficulties with infection control and accessing care. Many of these are reflected in the MadCovid project’s materials (https://madcovid.com/).

Telemedicine drew mixed views from staff; we would like to highlight some difficulties. Not everyone has a safe space to speak, may only have privacy in their bedroom or none at all. Telemedicine works better for those in better, not-overcrowded housing, so risks widening inequalities in access to care. For many of us, our home is our safety, and it is important to have distressing conversations elsewhere. Leaving the therapy room, we can leave some of our trauma behind. Videocalls may feel invasive - as though the clinician is in your bedroom - bringing up traumatic issues inside the home, where we cannot escape them. Any continuation of remote working will need to consider the safety implications of this, assessing its suitability for each individual.

It is vital that difficulty adhering to infection control guidance does not lead to blaming inpatients for viral spread. This is particularly important with restraint, where staff mentioned struggling to put on appropriate PPE in time to deal with an unfolding emergency. Wide area variations in restraint rates (https://www.mind.org.uk/media-a/4378/physical_restraint_final_web_version.pdf [17]; https://weareagenda.org/wp-content/uploads/2017/03/Restraint-FOI-research-briefing-FINAL1.pdf [18]), alongside personal experience, make us question whether restraint is ever truly unavoidable. If it places both staff and service users at risk of Covid-19 infection it is doubly dangerous. However challenging the situation, efforts must be renewed to reduce the iatrogenic distress, fear and anger which can lead to its use.

Historically slow-moving services have implemented change at breakneck speeds in response to this crisis despite significant difficulties. Service users have campaigned for changes for decades. It is time to implement these changes with the same urgency.

## Data Availability

The survey dataset is currently being used for additional research by the author research group and is therefore not currently available in a data repository. A copy of the survey is available at this web address: https://opinio.ucl.ac.uk/s?s=67819.

## Acknowledgments

We are very grateful to Seena Fazel (University of Oxford), Angela Hassiotis and Gill Livingston (both University College London) for their contributions to the development of survey questions in their specialist areas (forensic psychiatry, older people, and intellectual disability, respectively), and to Dr Lade Smith for her help in promoting the survey, especially by making a video with the Mental Elf encouraging staff from Black and minority ethnic groups to participate. We also thank the clinicians who took time to complete and feed back on our pilot survey, and all participants for taking time from the pressures of the pandemic to respond to the crisis.

## Declarations

### Ethics approval

The King’s College London research ethics committee approved this study (MRA-19/20-18372).

### Consent to participate

Information on participation was provided on the front page of the survey. By starting the survey, participants agreed that they had read and understood all this information.

### Consent for publication

It was explained on the front page of the survey that responses may be used in articles published in scientific journals and that these articles will not include any information which could be used to identify any participant.

### Code availability

Not applicable.

### Authors’ contributions

SJ, BLE, AS, SO, SC and SG made the initial plan for the study. All main authors contributed to protocol development. SJ drafted the survey, CDL developed the online version and all main authors commented. CDL and AS led on recruitment strategies. CDL and SL planned and conducted the statistical analysis. NVSJ, UF, AP and SO planned and conducted the qualitative analysis. RRO, TJ and PS coded and summarised qualitative data and contributed from a lived experience perspective, and RRO and TJ wrote the Lived Experience commentary. SJ, CDL and HK drafted and all authors contributed to and approved the final manuscript.

Group authors contributed to protocol development, commented on the survey, contributed to making and implementing recruitment plans, and coded and summarised qualitative data.

